# Risk of ESKD in related older living kidney donors

**DOI:** 10.1101/2022.03.11.22271853

**Authors:** Andrew Arking, Fawaz Al Ammary, Gabriella Kaddu, Allan B. Massie, Dorry L. Segev, Jacqueline Garonzik-Wang, Abimereki D. Muzaale

## Abstract

Living kidney donors who are biologically related to the recipient have higher risk for end-stage kidney disease (ESKD) compared with those who are unrelated to the recipient. This risk is greater for first-degree relatives than more distant relatives. To understand if this holds true for older donors, who were cleared for donation and might be past the peak-age for hereditary disease, we used donor data (SRTR) linked to ESKD registry data (CMS) and stratified donors by age (younger vs older [≥50 years]) and race (black, Hispanic, and white). Younger related donors of all racial groups had higher risk of ESKD compared with younger unrelated donors; however, only older related white and Hispanic donors had higher risk of ESRD compared with unrelated older donors (2.3-fold for white full-siblings and 1.9-fold for white parents/offspring; 3.3-fold for Hispanic full-siblings and 2.0-fold for Hispanic parents/offspring). Older related black donors did not have higher risk compared to older unrelated black donors (0.8-fold for black full-siblings and 0.5-fold for black parents/offspring). Our study points to an earlier age of onset of kidney disease in black donors with a family history of ESKD. Our findings call for programs that promote living donation among related older black donor candidates.

## INTRODUCTION

Donor-recipient relationship is linked to risk of ESKD in live kidney donors across various racial groups. The greatest risk is in identical twins, followed by those who donated to full-siblings, parents, and children; more distant relatives such as half-siblings have a slightly elevated risk for ESRD compared with unrelated donors. In this recent report, donor age and sex were treated as confounding characteristics; however, there was no attempt to investigate the discrete effects of age at donation on risk of ESKD [1].

The higher risk in related donors might be reflective of shared genetic risk between donor and recipient, and individuals with high-risk genotypes express kidney disease at a younger ages [2]. Given these factors, older related donors are less likely to possess a high-risk genotype because the phenotype would already be observable during the donor screening process, and such candidates would be deemed ineligible. For these outlined reasons, it might be necessary to separately infer the effect of donor-recipient relationship on risk of ESKD for younger donors and older donors [3].

To investigate whether the association between biological relationship and risk of ESKD would be distinct in the older donors (≥50 years) vs. their younger counterparts, we used national registry data as previously reported [1]. But in addition to stratifying all analyses and inferences by donor-recipient relationship and race, we further stratified by age at donation.

## METHODS

### Live Kidney Donors

This study used data from the Scientific Registry of Transplant Recipients (SRTR). The SRTR data system includes data on all donor, wait-listed candidates, and transplant recipients in the US, submitted by the members of the Organ Procurement and Transplantation Network (OPTN). The Health Resources and Services Administration (HRSA), U.S. Department of Health and Human Services provides oversight to the activities of the OPTN and SRTR contractors. This dataset has previously been described elsewhere [4]. This Institutional Review Board of Johns Hopkins University has determined that this study qualifies as exempt research under the DHHS regulations because it uses deidentified data.

Through this reporting, 149,205 adult live donors between October 1, 1987, and December 31, 2020, were included in this study (including those who donated to children). Asian donors were excluded from the study because our previous report demonstrated the effect of donor-recipient relationship on the donor’s risk for ESKD, but the sample size was too small for further stratification by age [1]. ESKD outcomes were ascertained by linkage to Centers for Medicare & Medicaid Services (CMS) medical evidence form 2728 (certification of ESKD; including records through August 30, 2020) using a combination of Social Security number, last name, first, middle name, or all 3; date of birth; and sex. ESKD was defined as the initiation of maintenance dialysis or receipt of a living or deceased donor kidney transplant, as previously reported. Donor-recipient relationship was ascertained by self-report at donor evaluation and was grouped as full sibling (including identical twins due to their small sample size), parent/offspring, half-sibling or other biological relative, and biologically unrelated (which included spouse and life partner). *A priori*, we decided to stratify age into two groups (<50 and ε50 years old), because we previously observed a direct association between age at donation and 15-year risk for ESKD, but only for those 50 years or older. For donors younger than 50 years, we previously observed an inverse association between age at donation and 15-year risk for ESKD: those 18 to 39 years old had a higher risk for ESKD compared with those aged 40 to 49 years [5].

### Cause of ESKD

As previously reported [1], we classified donor and recipient causes of ESKD into 8 broad categories: diabetes, hypertension or large-vessel disease, glomerulonephritis (GN), cystic kidney disease, other urologic disease, other cause, unknown cause, and missing cause. Diabetes includes type 2 (adult-onset type or unspecified type) and type 1 (juvenile type and ketosis-prone diabetes). Hypertension includes kidney disease caused by hypertension (no primary kidney disease), renal artery stenosis, renal artery occlusion, and cholesterol and renal emboli. GN includes GN (histology not examined), focal segmental GN, membranous nephropathy, membranoproliferative GN, dense deposit disease, immunoglobulin A (IgA) nephropathy, IgM nephropathy (proven by immunofluorescence), rapidly progressive GN, Goodpasture syndrome, postinfectious GN, and other proliferative GN. Cystic kidney disease included polycystic and medullary cystic kidney disease. All other diagnoses were grouped under other cause. Unknown cause was documented as such. Missing cause included all records with no information for cause of ESKD.

### Cumulative Incidence of ESKD

The outcome of interest was time to ESKD, for which time zero for all donors was the date of donation. Death before ESKD was a competing event. Cumulative incidence functions methods were used to estimate 20-year risk for ESKD, with a time scale of years since time zero.

### Adjusted Hazard Ratios of ESKD

Cause-specific hazards models were used to estimate multivariable-adjusted hazard ratios that accounted for age and sex by treating the competing events as censoring. Sensitivity analyses were performed treating recipient age and income as potential confounders. Because we were interested in investigating the familial basis of ESKD risk in donors, *a priori* we decided to perform these analyses in each racial group in addition to the two age groups. All analyses were performed using RStudio Version 1.3.959 for Windows (RStudio, PBC) running R 4.0.2. All hypothesis tests were 2 sided (α = 0.05).

## RESULTS

### Study Population

Among 149,205 live kidney donors, median age at donation was 41 years, 59% were female, 74% were white, 13% were Black, 13% were Hispanic. Seventy six percent of donors were younger than 50 and 24% were older than 50 years old. No donor had diabetes at baseline, but 3% had hypertension. Median eGFR for this population was 97 mL/min/ 1.73 m^2^, median systolic/diastolic blood pressure was 120/74 mm Hg, median body mass index was 27 kg/m^2^, 25% had a history of smoking cigarettes, 28% graduated from college, and 12% had postgraduate education. Regarding relationship to recipient, 36% were unrelated (including 11% who were spousal relations, but biologically unrelated), 8% were half-sibling or other biological relatives, 28% were parents/offspring, and 28% were full siblings. Older donors generally donated more recently than younger donors (average year of donation 2009 vs 2005) and to older recipients (average age 51 vs 43). Furthermore, older donors had six times the prevalence of hypertension at baseline (7.0% vs 1.2%). Characteristics varied by race and age group (Table 1).

**Table 1:** Characteristics of Study Population

|  | Black |  | Hispanic |  | White |  |
| --- | --- | --- | --- | --- | --- | --- |
|  | Younger<br>N=16486 | Older<br>N=2327 | Younger<br>N=17310 | Older<br>N=2803 | Younger<br>N=79374 | Older<br>N=30943 |
| Age, median years [IQR] | 34 [ 27- 40] | 54 [ 51- 58] | 34 [ 27- 41] | 54 [ 52- 58] | 38 [ 31- 44] | 55 [ 52- 59] |
| eGFR, ml/min/1.73m2 [IQR] | 109 [ 95-125] | 94 [ 82-109] | 109 [ 96-119] | 95 [ 83-101] | 99 [ 87-111] | 85 [ 75- 97] |
| Blood pressure, mmHg [IQR] |  |  |  |  |  |  |
| SBP | 120 [112-129] | 126 [118-135] | 118 [110-126] | 123 [115-133] | 120 [111-128] | 124 [115-134] |
| DBP | 74 [ 69- 80] | 78 [ 70- 82] | 72 [ 66- 79] | 74 [ 68- 80] | 74 [ 68- 80] | 75 [ 69- 81] |
| BMI, kg/m2 [IQR] | 28 [ 24- 31] | 28 [ 25- 30] | 27 [ 24- 30] | 27 [ 25- 30] | 26 [ 23- 30] | 26 [ 24- 29] |
| Year of Donation, median [IQR] | 2004 [1999-2011] | 2008 [2002-2014] | 2007 [2001-2013] | 2010 [2003-2015] | 2005 [1999-2011] | 2009 [2002-2015] |
| Recipient Age, median years [IQR] | 43 [ 31- 54] | 49 [ 33- 58] | 40 [ 28- 52] | 49 [ 32- 57] | 44 [ 32- 55] | 53 [ 39- 59] |
| Women, % | 57.2 | 62.6 | 57.8 | 65.3 | 58.9 | 64.0 |
| Diabetes, % | 0.0 | 0.0 | 0.0 | 0.0 | 0.0 | 0.0 |
| Hypertension, % | 1.2 | 6.7 | 0.7 | 5.3 | 1.4 | 7.8 |
| Smoke, % | 18.2 | 23.3 | 17.7 | 18.8 | 27.8 | 26.8 |
| Education, % |  |  |  |  |  |  |
| High School or Less | 34.8 | 30.8 | 50.2 | 49.6 | 28.6 | 25.2 |
| Attended College | 33.6 | 27.6 | 28.0 | 23.8 | 28.0 | 24.3 |
| Graduate | 23.0 | 27.0 | 17.2 | 17.9 | 31.3 | 31.5 |
| Postgraduate | 8.7 | 14.7 | 4.6 | 8.8 | 12.1 | 18.9 |
| Spouse or life partner, % | 7.9 | 18.6 | 9.5 | 20.7 | 8.5 | 19.3 |
| Donor-recipient relationship, % |  |  |  |  |  |  |
| Biologically unrelated to recipient | 22.0 | 35.6 | 26.0 | 38.1 | 35.8 | 51.7 |
| Half-sibling or other biological relationship | 11.0 | 7.7 | 8.7 | 5.1 | 7.8 | 5.5 |
| Parent/Offspring of recipient | 37.3 | 30.2 | 32.6 | 32.6 | 28.2 | 20.1 |
| Full-sibling of recipient | 29.7 | 26.5 | 32.7 | 24.1 | 28.3 | 22.7 |
| Recipient cause of ESKD, % |  |  |  |  |  |  |
| Diabetes | 15.5 | 15.8 | 15.1 | 16.9 | 14.7 | 15.1 |
| Hypertension or large vessel disease | 25.3 | 22.1 | 13.1 | 10.5 | 8.4 | 8.5 |
| Glomerulonephritis | 21.8 | 19.4 | 21.3 | 19.3 | 21.7 | 17.8 |
| Cystic kidney disease | 2.3 | 3.4 | 3.2 | 4.4 | 8.2 | 10.6 |
| Urological disease | 0.6 | 0.4 | 0.9 | 0.6 | 1.9 | 1.4 |
| Other Cause | 3.7 | 3.1 | 5.5 | 4.0 | 9.1 | 6.9 |
| Unknown Cause | 5.5 | 5.2 | 7.5 | 4.9 | 6.4 | 5.3 |
| Missing Cause | 25.3 | 30.4 | 33.3 | 39.6 | 29.6 | 34.3 |

### Cause of Kidney Failure

Recipient causes of kidney failure varied in relative frequency by race. For black donors, the recipient cause of kidney failure in both older and younger donors was most frequently hypertension (22% and 25% respectively), GN (19% and 22%), and diabetes (15% and 15%). For Hispanic donors, recipient cause of kidney failure was GN (19% and 21% for older and younger donors, respectively), diabetes (17% and 15%), and hypertension (10% and 13%). For white donors, the recipient cause of kidney failure was most frequently GN (18% and 22% in older and younger donors, respectively) and diabetes (15% and 15%), with cystic kidney disease (12%) in third place for older donors and hypertension in third for younger donors (8%).

### Cumulative Incidence of ESKD

Donors were followed up for a median of 12 (interquartile range, 6-18; maximum, 30) years and a total of 1,832,449 person-years, during which 432 donors developed ESKD. We estimated the incidence of ESKD according to age group, race, and donor-recipient biological relationship. For younger black donors, the 20-year risk for ESKD expressed per 10,000 donors was 174 (95% CI, 132-230) for full siblings, 106 (95% CI, 76-148) for parents/offspring, 58 (95% CI, 26-132) for half-sibling/other biological relatives, and 52 (95% CI, 16-167) for biologically unrelated donors. For older black donors, the 20-year risk for ESKD was 77 (95% CI, 18-325) for full siblings, 38 (95% CI, 5-266) for parents/offspring, 141 (95% CI, 20-958) for half-sibling/other biological relatives, and 93 (95% CI, 19-453) for biologically unrelated donors.

For younger Hispanic donors, the 20-year risk for ESKD was 30 (95% CI, 17-55) for full siblings, 25 (95% CI, 11-54) for parents/offspring, 34 (95% CI, 8-139) for half-sibling/other biological relatives, and 10 (95% CI, 1-72) for biologically unrelated donors. For older Hispanic donors, the 20-year risk for ESKD was 76 (95% CI, 18-324) for full siblings, 92 (95% CI, 33-252) for parents/offspring, 175 (95% CI, 25-1181) for half-sibling/other biological relatives, and 19 (95% CI, 3-131) for biologically unrelated donors. For younger white donors, the 20-year risk for ESKD was 27 (95% CI, 20-37) for full siblings, 30 (95% CI, 21-42) for parents/offspring, 11 (95% CI, 3-39) for half-sibling/other biological relatives, and 18 (95% CI, 9-37) for biologically unrelated donors. For older white donors the 20-year risk for ESKD was 84 (95% CI, 55-129) for full siblings, 70 (95% CI, 46-104) for parents/offspring, undefined for half-sibling/other biological relatives, and 23 (95% CI, 11-46) for biologically unrelated donors (Figure 1).

### Adjusted Risk for ESKD

We only adjusted for age and sex within each age stratum. Risk for ESKD as estimated by the Age- and Sex-adjusted hazard ratios varied by orders of magnitude across race. It remained consistent between age groups in white and Hispanic donors but differed between age groups in black donors. For related younger black donors, risk was 2.3-fold higher (95% CI, 1.2-4.5) for full siblings, 1.8-fold (95% CI, 0.9-3.6) for parents/offspring, and 1.3-fold (95% CI, 0.5-3.3) for half-sibling or other biological relatives compared with unrelated younger black donors; within the stratum, higher age within was associated with no higher risk: 0.8-fold (95% CI, 0.6-1.0). For related older black donors, risk was no higher compared to unrelated older black donors: 0.8-fold (95% CI, 0.1-5.5) for full siblings, 0.5-fold (95% CI, 0.1-3.5) for parents/offspring, and 1.9-fold (95% CI, 0.2-21.1) for half-sibling or other biological relatives; higher age within the stratum was similarly associated with no higher risk: 0.9-fold (95% CI, 0.4-9.3). For related younger Hispanic donors, risk was 1.5-fold higher (95% CI, 0.4-5.3) for full siblings, 1.6-fold (95% CI, 0.4-5.7) for parents/offspring, and 1.3-fold (95% CI, 0.2-7.6) for half-sibling or other biological relatives, but none of these differences reached statistical significance. For older related Hispanic donors, risk was 3.3-fold higher (95% CI, 0.3-33.6) for full siblings, 2.0-fold (95% CI, 0.2-18.4) for parents/offspring, and 5.0-fold (95% CI, 0.3-84.1) for half-sibling or other biological relatives, but none of these differences reached statistical significance. For related younger white donors, risk was 2.2-fold higher (95% CI, 1.3-3.9) for full siblings, 2.4-fold (95% CI, 1.4-4.2) for parents/offspring, but no higher for half-sibling or other biological relatives: 0.6-fold (95% CI, 0.2-2.2). For related older white donors, risk was 2.3-fold higher (95% CI, 1.3-4.3) for full siblings, 1.9-fold (95% CI, 1.0-3.5) for parents/offspring, and undefined for half-sibling or other biological relatives (Figure 2).

**Figure 2.**
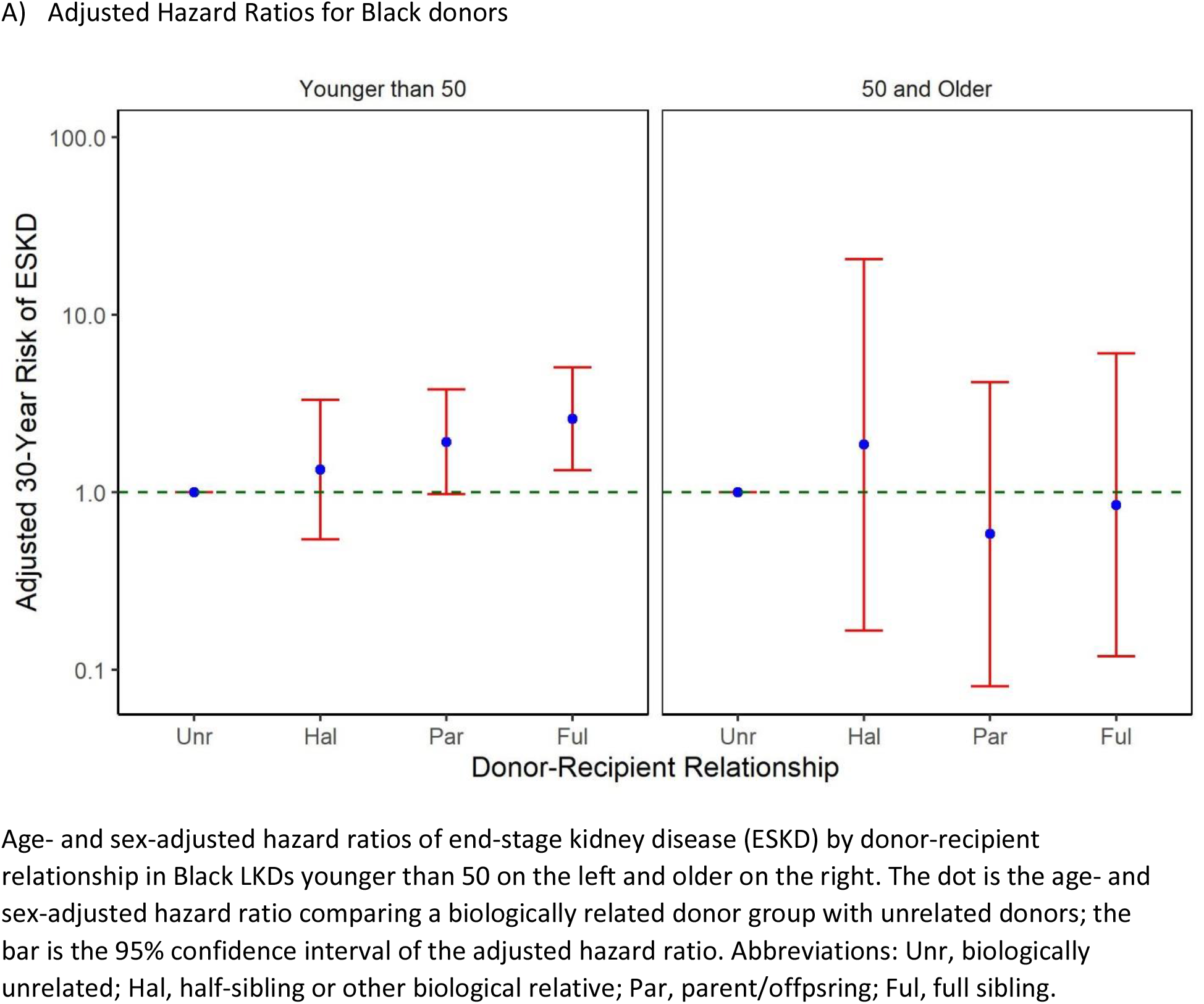
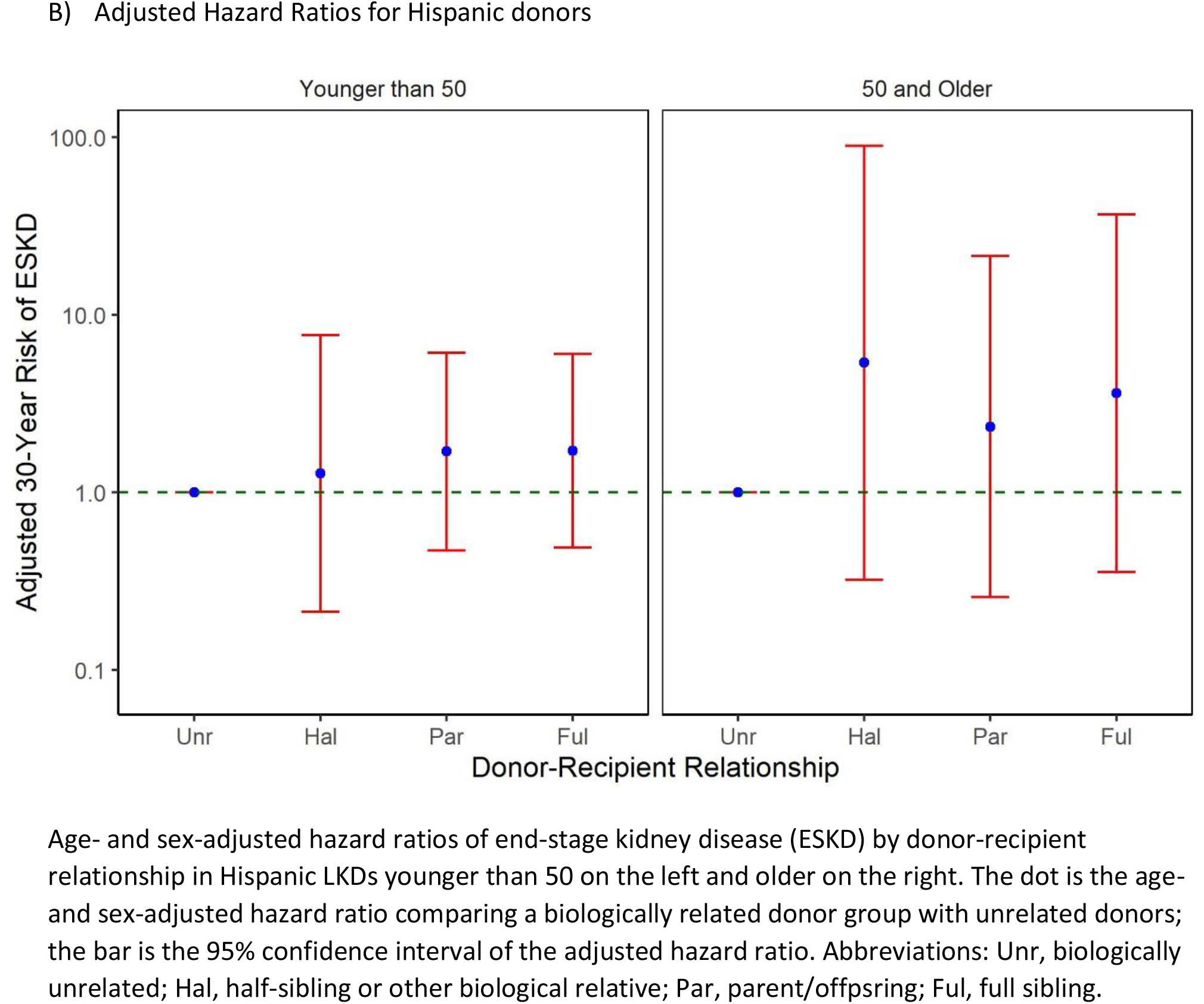
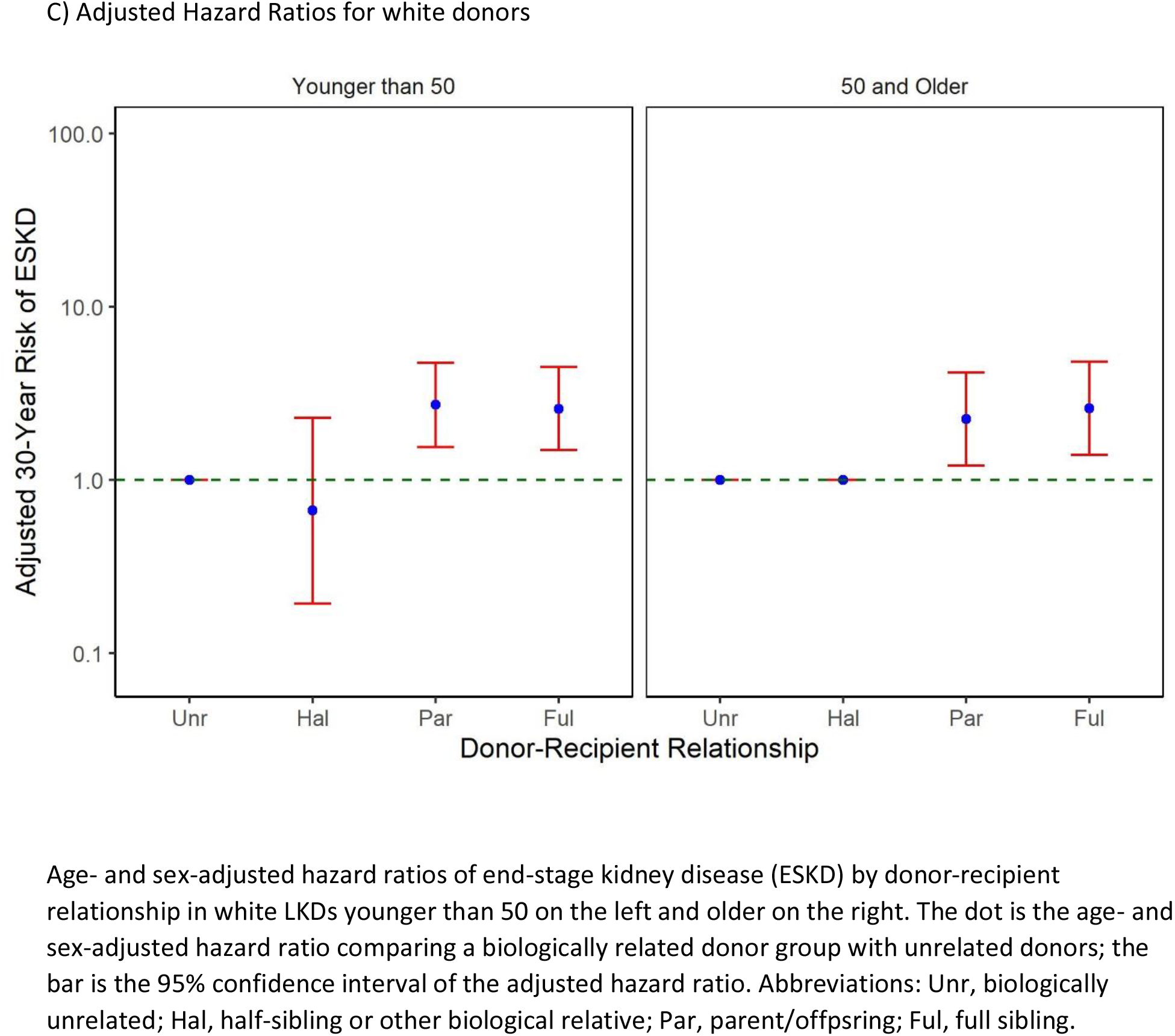

## DISCUSSION

In this national study, we observed elevated levels of risk for ESKD in living kidney donors based on biological relationship between donor and recipient across racial groups. Risk was generally higher for first-degree relatives (parent, offspring, and full-sibling) compared with more distant relatives, both of which exceeded the risk in unrelated donors. These trends persisted across racial groups, consistent with our previous report [6]. When donors were stratified by age (<50 vs. ε50 years old), related older and younger donors displayed elevated risks compared to unrelated older and younger donors for white and Hispanic donors; however, for black donors, related younger donors had elevated risk for ESKD while related older donors did not: full-siblings had a 2.3-fold risk in younger donors but no higher risk for older donors; parents/offspring had a 1.8-fold risk in younger donors but no higher risk in older donors. As such, for older black donors, biological relation did not indicate a higher risk of ESKD, unlike younger black donors and donors of all ages from other racial groups.

Unlike our previous study on risk of ESKD in biologically related donors, this study draws distinct inferences for younger vs. older black donors. This modification of the effect of donor-recipient relationship in black donors adds nuance to a previous observation that older age is not associated with risk of ESKD in black donors. Elevated risk of ESKD in related black donors stems from genetic and environmental risk factors shared by donor and recipient. In the general African American population, 13% of individuals carry two high-risk variants of apolipoprotein-L1 (APOL1), which is strongly associated with kidney disease [8, 9]. The average age at ESKD onset in patients with high-risk variants of APOL1 is 49 years old compared with 61 years for individuals with no high-risk variants [2]; the age of onset of earlier stages of chronic kidney disease might be 5-10 years earlier [10]. We might thus impute that donors older than 50 who have been screened for kidney disease and have been cleared for donation have a low likelihood of possessing the high-risk genotype, as it would likely manifest itself before 50. Similar patterns may be true for other genetic factors specific to Black donors and for environmental risk factors more common to this group. Related older black donors may thus be viewed as a highly selected population with no higher risk of ESKD compared with unrelated older black donors.

This study has two key strengths. First, no previous study of ESKD risk in living kidney donors stratified its analyses and inferences by degree of relatedness, race, and age. Stratifying by age allowed us to make novel inferences for related older black donors. Second, this study showed statistically significant higher risks for siblings and parents/offspring in white donors not found in previous analyses which used more relationship groups. That said, this study has several limitations as well. Grouping donors by age, race, and relationship leaves small sample sizes for each group and limits the statistical power for some analyses. Moreover, the rarity of ESRD in donors further limits the statistical power to examine several lifestyle-related risk factors beyond income that may affect first-degree relatives more than other relatives. Indeed, we did not have the statistical power to investigate sex as a biological factor contributing to some kidney diseases [11-13].

In conclusion, related donors have a higher risk of ESRD compared to unrelated donors across race and age groups. Black donors over 50 are an exception to this trend; biological relation is not associated with elevated ESRD risk, possibly because relevant genetic and environmental risks shared between relatives manifest themselves at a younger age in this population. As such, black donors older than 50 should not be discouraged from donating to relatives any more than non-relatives.

## Data Availability

All data produced in the present work are contained in the manuscript

## Abbreviations

(LKD): Living kidney donor
(ESKD): End-stage kidney disease

## ACKNOWLEDGEMENTS

This work was supported by grant number K24AI144954 (Segev) from the National Institute of Allergy and Infectious Diseases (NIAID) and K08AG065520-01 (Muzaale) from the National Institute on Aging. The analyses described here are the responsibility of the authors alone and do not necessarily reflect the views or policies of the Department of Health and Human Services, nor does mention of trade names, commercial products or organizations imply endorsement by the U.S. Government.

The data reported here have been supplied by the Hennepin Healthcare Research Institute (HHRI) as the contractor for the Scientific Registry of Transplant Recipients (SRTR). The interpretation and reporting of these data are the responsibility of the author(s) and in no way should be seen as an official policy of or interpretation by the SRTR or the U.S. Government.

